# Deficits in the knowledge of social norms and their underlying mechanisms in Alzheimer’s disease

**DOI:** 10.1101/2024.09.11.24312998

**Authors:** Thomas Carrier, Isabelle Rouleau, Marie-Anne St-Georges, Maxime Montembeault

## Abstract

**Background:** Compared to other components of social cognition, knowledge of social norms has received less attention, even more so in Alzheimer’s disease (AD). While semantic memory deficits have been identified early in the course of AD, no study has delved into the knowledge of social norms at these preliminary stages, although evidence suggests it shares common ground with semantic memory. In addition, it is unclear whether the knowledge of social norms in AD is associated socioemotional deficits, as seen in the behavioral variant of frontotemporal dementia (bvFTD). Finally, how social norms knowledge impairments predict behaviours in real-world settings remains unknown in the context of AD.

**Methods:** This study included 286 participants with mild cognitive impairment (MCI), 157 with AD, 285 with bvFTD along with 384 older healthy controls. All participants were selected from the National Alzheimer’s Coordinating Center. They completed the Social Norms Questionnaire, which assesses the tendency to break or overadhere to social norms. They also completed tests assessing executive, semantic and socioemotional functions, along with tests measuring spontaneous interpersonal behaviours.

**Results:** Between-group comparisons show that individuals with AD and MCI break and overadhere to social norms significantly more than HC, while they perform better than individuals with bvFTD. Knowledge of social norms was mainly associated with semantic knowledge across groups, and predicted insensitivity and disinhibition severity in patients.

**Conclusions:** This study suggests that declines in semantic memory likely play a key role in social norms knowledge decreases and that these decreases predict behavioural tendencies.

## Introduction

Worldwide, over 55 million individuals suffer from a major neurocognitive disorder, commonly known as dementia. Of these cases, 60% to 70% are due to Alzheimer’s disease (AD), making it the leading cause of major neurocognitive disorders ^1^. Impairment of episodic memory typically characterizes AD, but other deficits are often present ^2^. Indeed, many individuals suffering from AD exhibit deficits in linguistic, visuospatial, executive capacities, or behavior/personality ^3^. Consequently, McKhann et al. (2011) propose that an impairment in at least one other cognitive domain, aside from memory, is required for diagnosing AD ^3^.

Compared to other cognitive domains, behavior/personality, and more specifically social cognition, is relatively understudied and under-assessed in AD ^4–7^. It has been recently proposed that this underrepresentation is linked to the recent recognition of social cognition as a cognitive domain (newly included in the DSM-5) and inconsistencies in its definition ^5^. Social cognition refers to the cognitive processes that govern social interactions. It includes key aspects such as recognizing socially relevant information (e.g., interpreting others’ emotional expressions) as well as empathy, theory of mind and knowledge of social norms ^8–11^. Disruptions in these processes can affect an individual’s behavior and ability to interact socially and can negatively impact their interpersonal relationships ^4,12^.

An increasing body of research point to social cognition impairments in AD. Throughout the progression of AD, affected individuals may exhibit changes in social behavior, theory of mind, empathy, facial emotion recognition, and self-awareness ^4,12–15^. Furthermore, a recent systematic review highlighted a clear progression of social cognitive impairments from normal aging to AD, particularly affecting emotion recognition and theory of mind ^16^.

However, compared to other components of social cognition, knowledge of social norms has received less attention ^6^. Social norms are typically defined as the unwritten rules shared by members of a group or society that influence decision making by promoting or prohibiting behaviors ^17^. There are very few recognized assessment tools available to evaluate their knowledge ^18,19^. However, a new test was recently developed to evaluate knowledge of social norms in neurodegenerative diseases. This tool, called the Social Norms Questionnaire (SNQ), designed to determine how well individuals understand and accurately identify implicit, yet widely accepted, social boundaries in the predominant culture of the United States ^20^. It assesses the tendency to breach and overadhere to social norms.

Although relatively well studied in the behavioral variant of frontotemporal dementia (bvFTD), the SNQ has been used in a limited number of studies involving participants with AD. These studies have shown that individuals with AD are usually more likely to overadhere to social norms rather than breach them ^21,22^. Also, they have shown that the SNQ is valuable for distinguishing individuals with AD from those with bvFTD and individuals with psychiatric disorders, with deficits appearing less pronounced in AD than in people with bvFTD or psychiatric disorders ^21–23^.

Nonetheless, there are still many gaps to fill in our understanding of social norms knowledge in AD. Though semantic memory deficits have been identified early in the AD disease course (e.g., MCI), no study has yet examined knowledge of social norms in these prodromal stages ^24^. While most previous studies have focused on object or people knowledge, it would be interesting to investigate whether semantic deficits extend to the semantic category of social concepts in the MCI population and whether the SNQ can serve as an early cognitive marker of MCI. Furthermore, the cognitive mechanisms underlying the knowledge of social norms in AD are still poorly understood. Previous studies have shown that it might be associated with semantic memory and executive functions, but no study has investigated socioemotional abilities and/or loss of empathy as a contributing mechanism ^22,23^. It is unclear whether knowledge of social norms in AD is associated with deficits in social cognition, as it is likely to be in bvFTD. Finally, the ecological validity of the SNQ (i.e. how test performance predicts behaviours in real-world settings) remains unknown in the context of AD. In other words, it would be crucial to know whether deficits in social norms knowledge can predict altered spontaneous interpersonal behavior.

Therefore, this study aims to compare knowledge of social norms between healthy participants, individuals with MCI, patients diagnosed with AD, and patients with bvFTD. Participants with bvFTD were included as a comparison group to better understand the contribution of social cognition in AD-related deficits. We hypothesize that knowledge of social norms will be significantly lower in individuals with MCI compared to healthy individuals, significantly lower in individuals with AD compared to those with MCI and healthy individuals, and significantly lowest in patients with bvFTD. Subsequently, the study seeks to investigate the cognitive correlates of social norms knowledge in participants with MCI/AD and bvFTD. We hypothesize that knowledge of social norms will be mainly associated with semantic abilities in MCI/AD, whereas it will be mainly associated with socioemotional abilities in bvFTD. Finally, this study also aims to examine whether social norms knowledge predicts spontaneous interpersonal behaviors in AD and bvFTD. We hypothesize that social norms knowledge will significantly predict spontaneous interpersonal behaviors in AD and bvFTD.

## Methods

### Participants

This study includes 286 individuals with MCI, 157 with AD, 285 with bvFTD along with 384 cognitively unimpaired older healthy controls (HC). Participants were selected from the National Alzheimer’s Coordinating Center (NACC), which consists of data collected at Alzheimer’s Disease Research Centers (ADRC). This study used data from 21 ADRC for Uniform Data Set visits conducted between March 2012 and October 2022. Diagnoses were made by either a consensus team or a single physician according to relevant current diagnostic criteria at the time of testing ^3,25,26^. To be considered healthy, HC had to be cognitively intact, as indicated by CDR® Dementia Staging Instrument score of 0. Additionally, all non-AD participants were required to be older than 47 years old, which corresponds to the age of the youngest AD participant. Written informed consent is obtained from all participants by all ADRCs. Approvals from the Institutional Review Boards of all participating institutions were obtained by the NACC.

## Measures

### Social Norms Questionnaire (SNQ)

The SNQ measures the degree to which a person understands and can accurately identify implicit but widely accepted social boundaries in the dominant United States culture ^20^. It consists of 22 Yes-No questions to be completed by the subject in the presence of a qualified psychologist or psychometrist during face-to-face testing. Subjects are instructed to determine whether a behavior would be appropriate in the presence of an acquaintance (not a close friend or family member) according to “mainstream” culture. A total score can be calculated (ranging from 0 to 22), with higher score indicating greater knowledge of social norms. Additionally, the tendency to breach social norms (Break score, ranging from 0 to 12) and to adhere to norms excessively (Overadhere score, ranging from 0 to 10), which acts as a control scale, can be calculated.

### Other measures

See Supplementary Material for details.

### Disease severity

To describe the sample and measure disease severity, the CDR® Dementia Staging Instrument (CDR®) and the Montreal Cognitive Assessment (MoCA) were used to assess disease severity ^27,28^.

### Semantic Memory

To assess semantic memory, the Semantic Associates Test and the Multilingual Naming Test (MINT) were used ^29,30^.

### Executive functions

To assess executive functions, the part B of the Trail Making Test (TMT) and the Backward Number Span Test were used ^31,32^.

### Socioemotional abilities

To assess socioemotional abilities, the Interpersonal Reactivity Index (IRI) and the Revised Self-Monitoring Scale (RSMS) were used ^33,34^.

### Spontaneous interpersonal behaviours

To measure real-life interpersonal behaviors, the Social Behavior Observer Checklist (SBOCL) and the Disinhibition scale of the Neuropsychiatric Inventory-Questionnaire (NPI-Q) were used ^35,36^.

### Statistical Analysis

Data were analyzed using R for Windows version 4.4.1. ANOVAs were performed to assess groups differences on demographic measures and overall cognitive performance, followed by Tukey HSD’s post-hoc test. Chi-squared test was used to assess group differences on dichotomous variables (i.e., sex). Between-group differences in the SNQ total score, Overadhere errors and Break scores were examined using ANCOVAs adjusted for participants’ age, education, and sex. Pairwise comparisons were conducted with Tukey-Kramer’s post-hoc tests. Furthermore, partial spearman correlations, adjusted for sex, age, education, and disease severity as measured with CDR®’s sum of boxes score, were conducted between SNQ variables and 1) semantic memory, as measured by the Semantic Associates Test and the MINT; 2) executive performance, evaluated using the Backward Number Span and the TMT; and 3) behavioral impairments, as indicated by scores on the IRI and the RSMS. Groups of MCI and AD participants were combined as both were considered on the AD spectrum. Finally, to assess whether social norms knowledge deficits predict interpersonal behaviors in AD and bvFTD, analyses of regression were conducted between SNQ break score, NPI-Q disinhibition severity, and SBOCL insensitivity scale, while controlling for participants age, education, sex, and disease severity. As in the previous analyses, groups of MCI and AD participants were combined

## Results

### Data preparation

Social norms questionnaire response ratios of Yes/No responses per participant were inspected to exclude participants with response bias great enough to render score meaningless (e.g., answering Yes or No to almost all, or all, items of the SNQ). That is, according to the SNQ’s scoring instructions, cases with scores greater than or equal to 5, or is less than 0.3 were excluded (15 HC, 28 MCI, 38 AD and 107 bvFTD) ^9^.

### Demographic Data

The analyses included 286 individuals with MCI, 157 with AD, 285 with bvFTD along with 384 HC. Demographic data is presented in Table 1. Participants in the MCI and bvFTD group were significantly older than participants in the HC and AD group (F(3, 1108) = 19.9, *p* < .001, η^2^ = 0.05). The percentage of women was significantly higher in the HC group than in the bvFTD group ( χ^2^(3) = 26.2, *p* < .001), but equivalent in all other groups. There were no significant differences between groups in terms of level of education (*p* = .39). Participants with bvFTD had significantly higher CDR® sum of boxes score than AD participants, who had significantly higher score than MCI participants, who had significantly higher score than HC participants (F(3, 1108) = 562.2, *p* < .001, η^2^ = 0.60). The same differences were observed between groups in MoCA total scores (F(3, 752) = 159.7, *p* < .001, η^2^ = 0.39). Therefore, we controlled for demographic variables and disease severity in all analyses.

**Table 1.** Participants Characteristics According to Group.

|  | HC | MCI | AD | bvFTD |
| --- | --- | --- | --- | --- |
| n | 384 | 286 | 157 | 285 |
| Age | 60.5 (9.4)<sup>a,c</sup> | 64.6 (8.7)<sup>b,d</sup> | 61.1 (6.7)<sup>a,c</sup> | 64.6 (8.1)<sup>b,d</sup> |
| Sex, % women | 57.6<sup>c</sup> | 48.6 | 49.0 | 37.5<sup>d</sup> |
| Education | 16.0 (2.5) | 16.1 (2.7) | 15.7 (2.9) | 15.7 (2.9) |
| CDR®, sum of boxes | 0.0 (0.0)<sup>a,b,c</sup> | 1.9 (1.5)<sup>b,c,d</sup> | 4.8 (2.5)<sup>a,c,d</sup> | 6.1 (3.2)<sup>a,b,d</sup> |
| MoCA, total score | 27.2 (2.4)<sup>a,b,c</sup> | 22.0 (4.7)<sup>b,c,d</sup> | 16.5 (5.7)<sup>a,c,d</sup> | 19.8 (6.0)<sup>a,b,d</sup> |

*Note* . The table presents mean (SD) values, unless indicated otherwise. For all comparisons, statistical significance criterion was $p < .05$ .
Abbreviations: HC, healthy controls; MCI, mild cognitive impairment; AD, Alzheimer's disease; bvFTD, behavioural variant of frontotemporal dementia; MoCA, Montreal Cognitive Assessment.
<sup>a</sup> $p < .05$ with MCI.
<sup>b</sup> $p < .05$ with AD.
<sup>c</sup> $p < .05$ with bvFTD.
<sup>d</sup> $p < .05$ with HC.

### Between-Group Analysis

Analyses of variance adjusted for age, education and sex revealed significant main effects of participant’s diagnosis on SNQ Total score (F(3, 1087) = 66.7 , *p* < .0001, η_p_^2^= 0.16), Break score (F(3, 1087) = 44.6 , *p* < .0001, η_p_^2^ = 0.11) and Overadhere score (F(3, 1087) = 32.9 , *p* < .0001, η_p_^2^= 0.08). Detailed results are presented in Table 2 and Figure 1. Tukey-Kramer post hoc tests indicated that the mean Total scores of the MCI, AD and bvFTD groups were significantly lower than the HC group. Participants with bvFTD had lower mean Total scores than the MCI and AD groups. The mean Break and Overadhere scores of the MCI, AD and bvFTD groups were significantly higher than the score of the HC group. Participants with bvFTD had higher mean Break scores than the MCI and AD groups, and higher mean Overadhere scores than the MCI group. The SNQ Overadhere score did not differ significantly differ between the AD and bvFTD groups (*p* = .10). The SNQ mean Total, Break and Overadhere scores did not differ significantly between the AD and MCI groups (*p* = .74; *p* = .92; *p* = .82).

**Figure 1:**
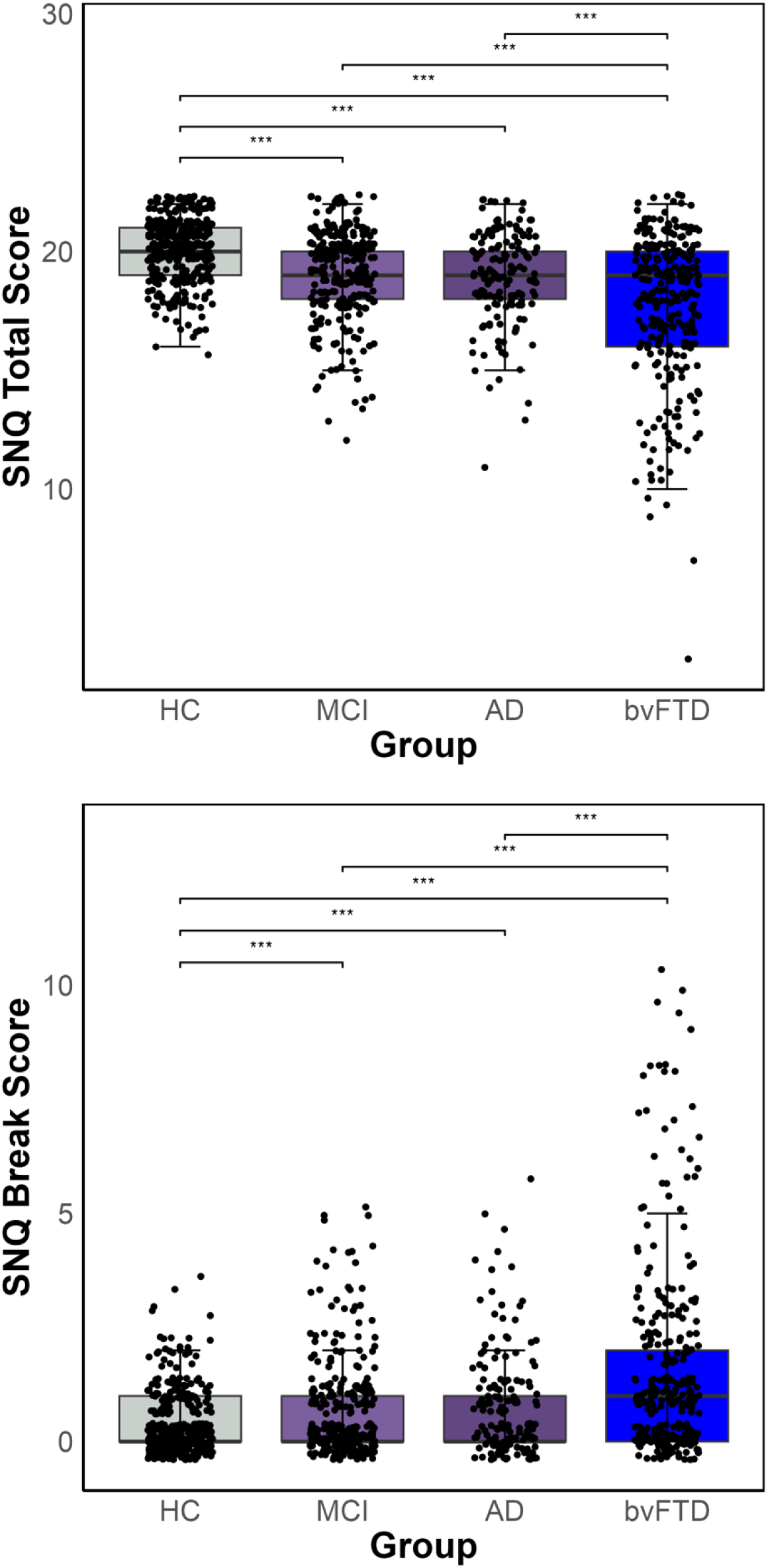
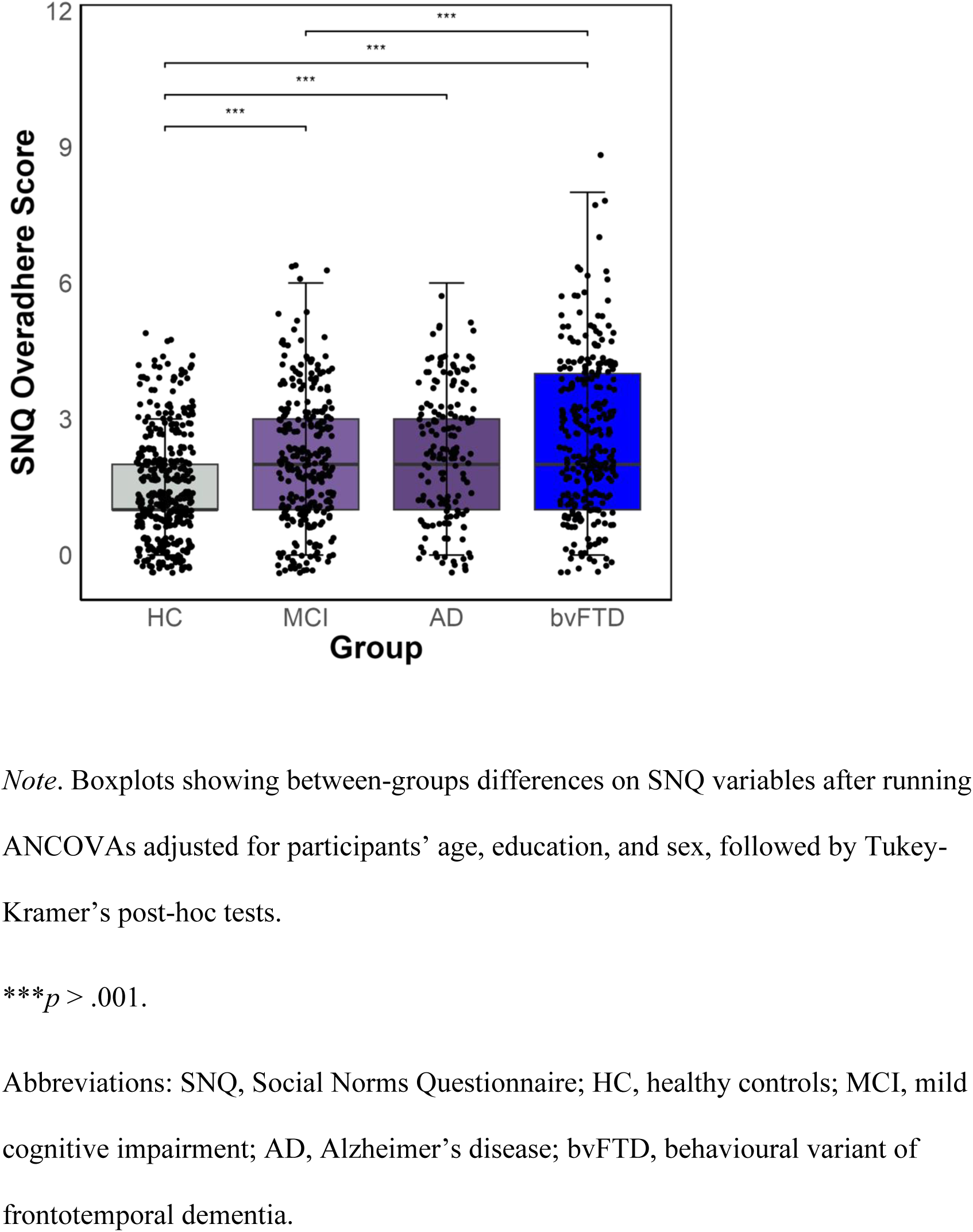
Group Differences in SNQ Total, Break and Overadhere Scores

**Table 2.** SNQ Performance Across Groups.

|  | Mean |  |  |  |
| --- | --- | --- | --- | --- |
|  | HC | MCI | AD | bvFTD |
| Total score | 20.1 (1.3)<sup>a,b,c</sup> | 19.1 (1.9)<sup>c,d</sup> | 18.9 (1.9)<sup>c,d</sup> | 17.7 (3.1)<sup>a,b,d</sup> |
| Break score | 0.4 (0.7)<sup>a,b,c</sup> | 0.8 (1.1)<sup>c,d</sup> | 0.9 (1.2)<sup>c,d</sup> | 1.7 (2.2) |
| Overadhere score | 1.5 (1.2)<sup>a,b,c</sup> | 2.1 (1.4)<sup>c,d</sup> | 2.3 (1.5)<sup>d</sup> | 2.6 (1.7)<sup>a,d</sup> |

Note. The table presents mean (SD) values. For all comparisons, statistical significance criterion was $p < .05$ .
Abbreviations: HC, healthy controls; MCI, mild cognitive impairment; AD, Alzheimer's disease; bvFTD, behavioural variant of frontotemporal dementia; MoCA, Montreal Cognitive Assessment.
<sup>a</sup> $p < .05$ with MCI.
<sup>b</sup> $p < .05$ with AD.
<sup>c</sup> $p < .05$ with bvFTD.
<sup>d</sup> $p < .05$ with HC.

### SNQ Cognitive Correlates

#### Semantic memory

Detailed results of norm transgression and overadherance correlates are respectively presented in Figure 2 and Figure 3. In the MCI/AD and bvFTD groups, partial spearman correlations, controlling for age, education, sex, and disease severity revealed that semantic memory as measured by the Semantic Associates Test and the MINT was significantly associated with norm transgression (i.e., SNQ break score) and norm overadherance (i.e., SNQ overadhere score). In the total sample (HC, MCI, AD and bvFTD), norm transgression was significantly associated with performance on the Semantic Associates Test and the MINT. Additionally, norm overadherance was significantly associated with performance on the MINT.

**Figure 2:**
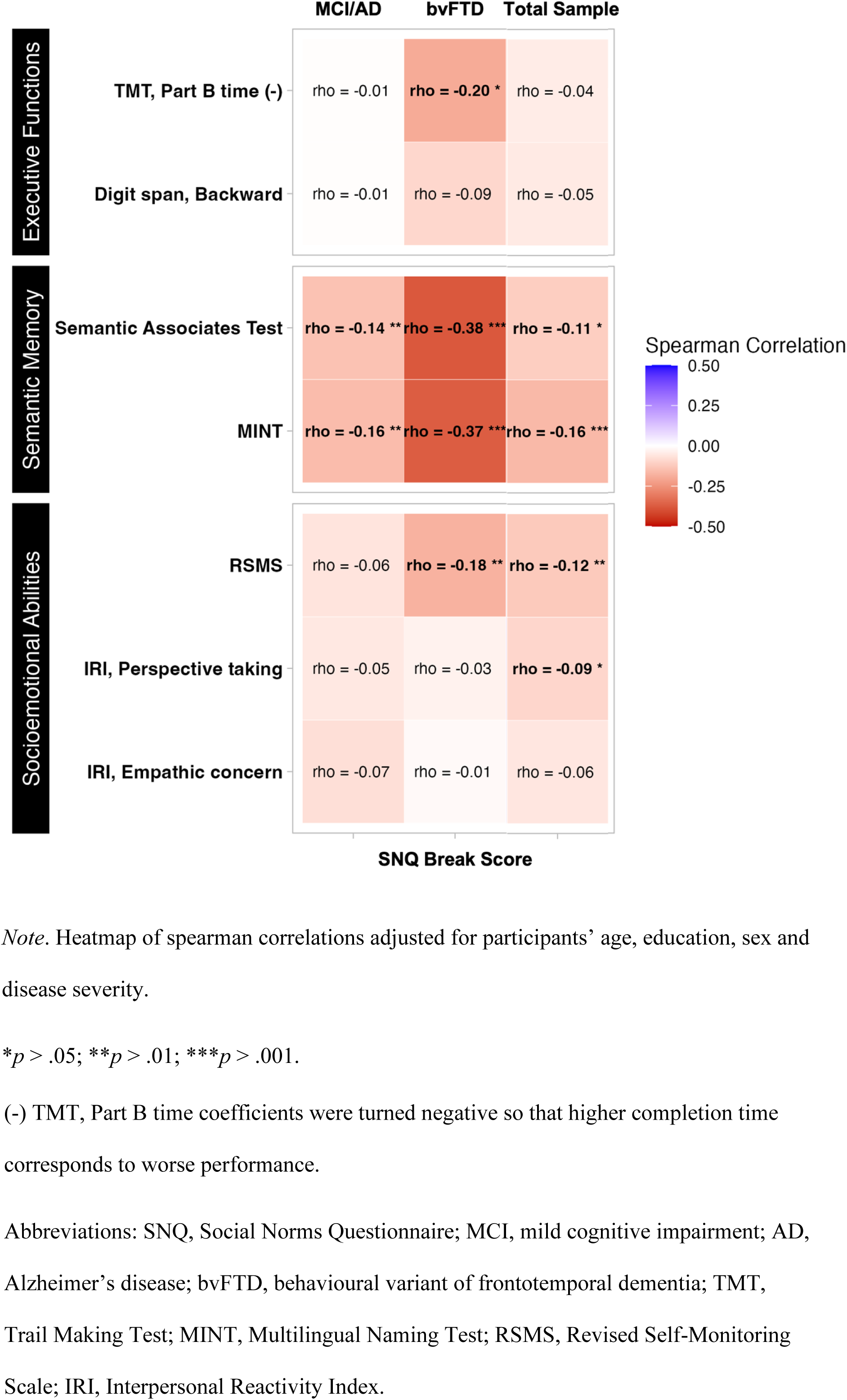
Norm Transgression Correlates in MCI/AD, bvFTD and in the Total Sample

**Figure 3:**
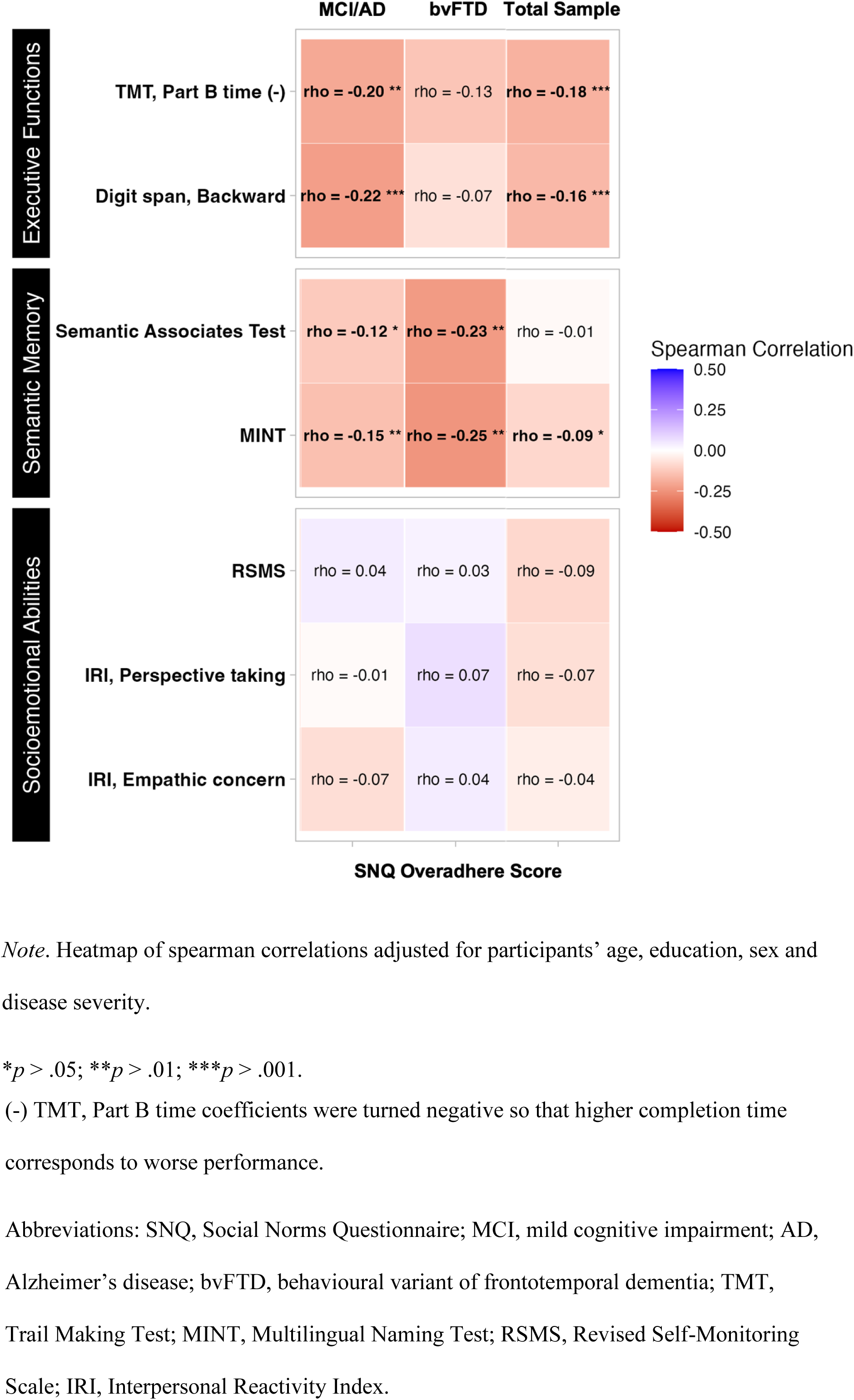
Norm Overadherance Correlates in MCI/AD, bvFTD and in the Total Sample

### Executive function

In MCI/AD participants, norm overadherance was significantly associated with the TMT part B completion time and with the backward digit span. In bvFTD participants, norm transgression was significantly associated with the TMT part B completion time. In the total sample, norm overadherance was significantly associated with the TMT part B completion time and with the backward digit span.

### Socioemotional abilities

In MCI/AD participants, SNQ performance was not associated with socioemotional abilities. In bvFTD, norm transgression was significantly correlated with score on the RSMS. In the total sample, norm transgression was significantly associated with score on the RSMS and on the perspective taking scale of the IRI.

### SNQ-based predictions of spontaneous interpersonal behaviours

In the total patient’s sample (i.e., MCI, AD and bvFTD), controlling for participants’ age, education, sex, and disease severity, norm transgression significantly predicted insensitivity on the SBOCL (*β* = 0.06, *p* < .01; *R^2^_adj_* (717) = 0.07, *p* < .001) and disinhibition severity (*β* = 0.06, *p* < .05; *R^2^_adj_* (696) = 0.15, *p* < .001). In MCI/AD only, norm transgression on the SNQ was a significant predictor of SBOCL insensitivity cluster (*β* = 0.10, *p* < .001; *R^2^_adj_* (434) = 0.04, *p* < .001), but not of disinhibition severity on the NPI. In bvFTD only, norm transgression did not significantly predict insensitivity nor disinhibition severity (*p* > .05). All regression plots are presented in Figure 4.

**Figure 4.**
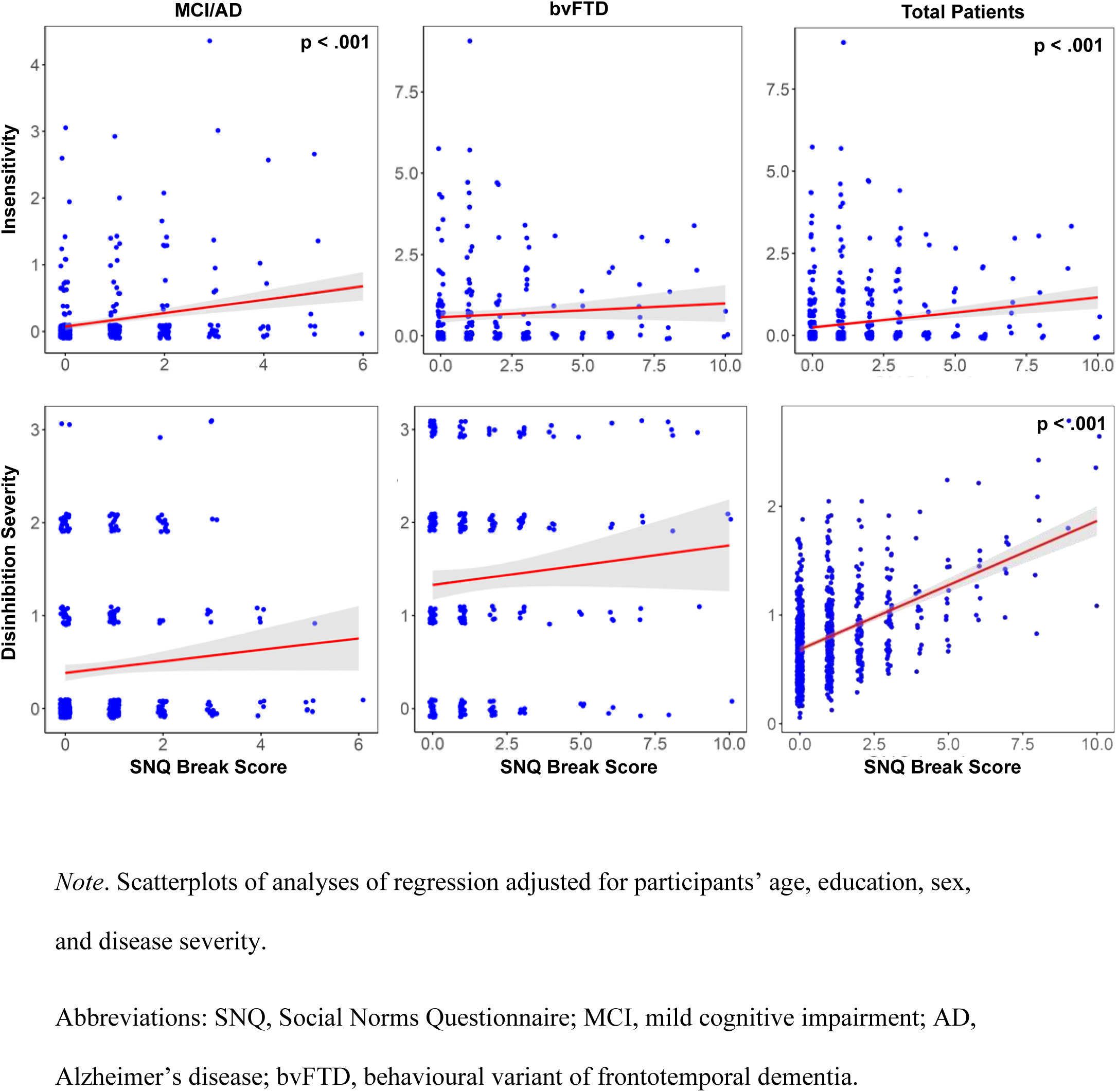
Relationship Between Norm Transgression and Insensitivity Across Samples

## Discussion

This study aimed to 1) compare the knowledge of social norms between healthy participants, individuals with MCI, those diagnosed with AD and patients with bvFTD, 2) investigate the cognitive correlates of knowledge social norms in patients with MCI/AD and bvFTD and 3) to investigate whether social norms knowledge predicts spontaneous interpersonal behaviors in MCI/AD and bvFTD. First, results suggest that alterations in social norms knowledge are evident in AD and bvFTD, but also in MCI, such that they are more likely to breach social norms and overadhere to them. Second, results show social norms transgression was associated only with semantic memory in MCI/AD, whereas it was additionally associated with executive function and empathy in bvFTD. Social norms overadherance was associated with executive function and semantic memory in MCI/AD, while it was solely associated with semantic memory in bvFTD. Third, norm transgression significantly predicted insensitivity during cognitive testing and disinhibition severity in daily life in the total patient sample, as well as insensitivity in MCI/AD patients only.

Consistent with previous studies observing decline in semantic memory in MCI ^24^, decline in social norms knowledge were observed in MCI. Indeed, participants suffering with MCI were more likely to breach and overadhere to social norms than HC participants. These decline in social norms knowledge were as severe as those observed in AD. Although AD participants included in the study had mild dementia, these results still highlight the fact that the SNQ may be clinically useful even in early stages of the AD spectrum. To our knowledge, this is the first study investigating social norms knowledge in MCI. Consistent with previous studies, AD patients had better social norms knowledge than bvFTD patients ^21–23,37,38^. This suggests that the SNQ may not only be an early marker for the diagnosis of patients on the AD spectrum but may also aid in the differential diagnosis between AD and bvFTD. Interestingly, while initial research highlighted the importance of episodic memory and executive tests for the differential diagnosis of AD and bvFTD, it is becoming increasingly clear that social cognition tests may be more appropriate for this clinical purpose ^39^.

While decline in knowledge of social norms has been found in all clinical groups, it is possible that similar deficits are caused by different underlying cognitive mechanisms. Failing an item on the SNQ could be due to different reasons. For example, it could be due to loss of semantic knowledge underlying social norms, executive dysfunction (e.g., perseveration, rigidity, etc.) and/or lack of socioemotional abilities guiding social behaviour (e.g., empathy).

Our results suggest that in AD, norm transgression was only significantly associated with semantic memory. However, in bvFTD, norm transgression was also associated with cognitive flexibility, empathy, in addition to semantic memory. This is coherent with common symptoms of bvFTD, such as executive dysfunction and loss of empathy ^40^. When norm transgression correlations were evaluated in the total sample, it was significantly associated with semantic memory and empathy. Taken together, these findings suggest that social norms knowledge is mostly associated with semantic memory, which is unsurprising since knowledge of social norms is a form of semantic knowledge. Additionally, recent studies have highlighted that social knowledge is processed by the same brain areas and networks implicated in non-social or object knowledge. More specifically, while numerous studies have demonstrated the core importance of the anterior temporal lobes (ATLs) in the storage and processing of conceptual knowledge ^41^, a growing body of evidence reveals broader cognitive implications of the ATLs. In fact, several studies suggest that social semantics are maintained by a distributed set of cerebral regions with the ATLs at their core ^42–44^. In this context, the ATLs are thought to be semantic knowledge hubs that interact with multiple brain regions to enable appropriate social behaviour ^45^. For example, in a previous study using the SNQ with AD and bvFTD patients, lower volumes of the ATLs were found to be significantly correlated with higher norm transgressions ^23^. Moreover, Rouse et al. (2023)^46^ showed that both social and non-social types of semantic knowledge impairments were associated with bilateral ATLs atrophy in FTD participants. Altogether, these findings suggest that impairments in social norms knowledge may be driven primarily by semantic disturbances.

The role of social norms knowledge in predicting spontaneous interpersonal behavior has been evaluated to better understand the life impact of decreases in social knowledge. In fact, the lack of ecological validity of social cognition tests has been criticized and may contribute to the fact that social cognition is not routinely assessed in a clinical setting ^47,48^. Our results suggest that, in the total patient sample, social norms performance predicts insensitivity during the cognitive assessment along with the severity of disinhibition over the previous month, as reported by an informant. Given that insensitivity encompasses a range of behaviors as evaluated in the SBOCL, our results suggest that norm transgression is associated with behaviors such as “too little self-consciousness”, “insensitive to other’s embarrassment”, “overly disclosing/ inappropriately familiar”. In the NPI, disinhibition was defined as acting impulsively, for instance, talking to strangers as if they were acquaintances or making remarks that could hurt others’ feelings. Although insensitive and disinhibited behaviors are most often seen in individuals suffering from bvFTD when compared to AD individuals and that social norm knowledge is generally more impaired in bvFTD than it is in AD, these behaviors were not predicted by social norms knowledge in bvFTD. This unexpected result might be due to methodological factors such as the absence of formal and specific assessment of socially inappropriate behavior. Alternatively, this result may suggest that socially inappropriate behaviors are driven by other deficits than loss of social semantic knowledge (e.g., empathy or cognitive disinhibition). Future studies with proper assessments of these constructs could clarify the strongest predictors of daily socially inappropriate behaviors in bvFTD. Surprisingly, significant association between insensitivity and norm transgression was observed in AD. Taken together, we believe these results provide support for the ecological validity of the SNQ.

This study has some limitations. Diagnoses of dementia were clinical. Therefore, they were not supported by biomarkers evidence. Additionally, the sample of AD participants included in this study was relatively young compared to MCI and bvFTD participants, which is atypical in community-based patients. Thus, generalizability of results may be affected.

In conclusion, this study suggests that declines in semantic memory likely play a key role in the decline in social norms knowledge in both AD and bvFTD. In everyday settings, these declines appear to be associated with insensitivity and the severity of disinhibited behaviors.

Overall, this study extends the findings of previous studies by focusing on the underlying mechanisms of social norms knowledge. This study may help clinicians and researchers better understand the potential behavioral consequences of specific cognitive impairments, such as semantic memory deficits. Future research should assess the progression of knowledge of social norms impairments over the course of AD and bvFTD, while delineating their cognitive and cortical associations. This may reveal key mechanisms driving behavioural dysfunction and thus contribute to the development of effective behavioral interventions.

## Data availability

The data that support the findings of this study are available from the National Alzheimer’s Coordinating Center (NACC) database. Restrictions apply to the availability of these data, which were used under license for this study. Data are available at https://naccdata.org/requesting-data/submit-data-request with the permission of the NACC.

## Data Availability

All data produced in the present study are available upon reasonable request to the authors

## Acknowledgments

The NACC database is funded by NIA/NIH Grant U24 AG072122. NACC data are contributed by the NIA-funded ADRCs: P30 AG062429 (PI James Brewer, MD, PhD), P30 AG066468 (PI Oscar Lopez, MD), P30 AG062421 (PI Bradley Hyman, MD, PhD), P30 AG066509 (PI Thomas Grabowski, MD), P30 AG066514 (PI Mary Sano, PhD), P30 AG066530 (PI Helena Chui, MD), P30 AG066507 (PI Marilyn Albert, PhD), P30 AG066444 (PI John Morris, MD), P30 AG066518 (PI Jeffrey Kaye, MD), P30 AG066512 (PI Thomas Wisniewski, MD), P30 AG066462 (PI Scott Small, MD), P30 AG072979 (PI David Wolk, MD), P30 AG072972 (PI Charles DeCarli, MD), P30 AG072976 (PI Andrew Saykin, PsyD), P30 AG072975 (PI David Bennett, MD), P30 AG072978 (PI Neil Kowall, MD), P30 AG072977 (PI Robert Vassar, PhD), P30 AG066519 (PI Frank LaFerla, PhD), P30 AG062677 (PI Ronald Petersen, MD, PhD), P30 AG079280 (PI Eric Reiman, MD), P30 AG062422 (PI Gil Rabinovici, MD), P30 AG066511 (PI Allan Levey, MD, PhD), P30 AG072946 (PI Linda Van Eldik, PhD), P30 AG062715 (PI Sanjay Asthana, MD, FRCP), P30 AG072973 (PI Russell Swerdlow, MD), P30 AG066506 (PI Todd Golde, MD, PhD), P30 AG066508 (PI Stephen Strittmatter, MD, PhD), P30 AG066515 (PI Victor Henderson, MD, MS), P30 AG072947 (PI Suzanne Craft, PhD), P30 AG072931 (PI Henry Paulson, MD, PhD), P30 AG066546 (PI Sudha Seshadri, MD), P20 AG068024 (PI Erik Roberson, MD, PhD), P20 AG068053 (PI Justin Miller, PhD), P20 AG068077 (PI Gary Rosenberg, MD), P20 AG068082 (PI Angela Jefferson, PhD), P30 AG072958 (PI Heather Whitson, MD), P30 AG072959 (PI James Leverenz, MD).

