## Supplementary Material for "Deficits in the knowledge of social norms and their underlying mechanisms in Alzheimer’s disease"

### Measures

**Semantic Memory**. The Semantic Associates Test is a test of knowledge of the meaning of objects in which subjects are asked to pick one of two pairs of objects that "go together" [1]. Scores range from 0 to 16. Higher scores are interpreted as reflecting better semantic memory. The MINT is a test of visual object naming where line drawings are presented to the subjects with the instruction to name of the object [2]. Scores range from 0 to 32. Higher scores are interpreted as reflecting better semantic memory.

**Executive functions**. The TMT part B is a test of processing speed and cognitive flexibility [3]. The subject must connect 25 circles while alternating between numbers and letters in an ascending order (e.g., A to 1; 1 to B; B to 2; 2 to C). The subject's performance is judged in terms of the time, in seconds, required to complete each the trial. Higher time of completion is interpreted as reflecting slower processing speed and/or poorer cognitive flexibility. The Backward Number Span Test measures working memory [4]. After being read numbers aloud, with sequences ranging from 2 to 9 numbers, the subjects are instructed to repeat the numbers in reverse order, or backward. The total number of correct trials (0-14) were used to judge the participants' performance. Higher scores are interpreted as having a higher working memory capacity.

**Socioemotional abilities**.The IRI is completed by an informant and measures empathy in everyday social interactions [5]. The informant is asked to indicate how well statements describe the subject's current typical behavior. The IRI is composed of 2 seven-item scales: Empathic Concern and Perspective-taking. The IRI performance is assessed with theses scales, ranging from 7 to 35 each. Higher scores are interpreted as reflecting a greater degree of empathy. The RSMS is completed by an informant and is designed to assess the degree to which subjects attend to others' socioemotional signals and allow those signals to influence their behavior [6]. The informant is asked to indicate how well statements describe the subject's current typical behavior. The RSMS is composed of 13 items, yielding a total score ranging from 0 to 65. Higher scores are interpreted as reflecting a greater degree of interpersonal sensitivity and responsiveness.

**Spontaneous interpersonal behaviours**. The SBOCL is utilized by clinicians to evaluate the severity and frequency of specific behaviors observed during a cognitive assessment session [7]. These behaviors are subdivided into multiple descriptors and 3 symptom clusters can be derived from the descriptors. These clusters are (1) Disorganized, (2) Reactive, and (3) Insensitive. The later cluster was included in the present study. Higher scores are interpreted as reflecting a greater degree of behavioral disturbance. The NPI-Q is a self-administered questionnaire filled out by informants about the patients they care for [8]. Presence and severity of neuropsychiatric symptoms are assessed. Data related to disinhibition was used in the present study.

2. Gollan TH, Weissberger GH, Runnqvist E, Montoya RI, Cera CM. Self-ratings of spoken language dominance: A Multilingual Naming Test (MINT) and preliminary norms

for young and aging Spanish–English bilinguals. *Bilingualism: language and cognition*. 2012;15(3):594-615.

3. Reitan RM. Validity of the Trail Making Test as an indicator of organic brain damage. *Perceptual and motor skills*. 1958;8(3):271-276.

4. Weintraub S, Besser L, Dodge HH, et al. Version 3 of the Alzheimer Disease Centers' Neuropsychological Test Battery in the Uniform Data Set (UDS). *Alzheimer Dis Assoc Disord*. Jan-Mar 2018;32(1):10-17. doi:10.1097/wad.0000000000000223

5. Davis MH. A multidimensional approach to individual differences in empathy. 1980;

6. Lennox RD, Wolfe RN. Revision of the self-monitoring scale. *J Pers Soc Psychol*. Jun 1984;46(6):1349-64. doi:10.1037//0022-3514.46.6.1349

7. Rankin KP, Toller G, Gavron L, et al. Social Behavior Observer Checklist: Patterns of Spontaneous Behaviors Differentiate Patients With Neurodegenerative Disease From Healthy Older Adults. *Front Neurol*. 2021;12:683162. doi:10.3389/fneur.2021.683162

8. Cummings J. The Neuropsychiatric Inventory: Development and Applications. *J Geriatr Psychiatry Neurol*. Mar 2020;33(2):73-84. doi:10.1177/0891988719882102
